# Multiplexed Detection and Quantification of Human Antibody Response to COVID-19 Infection Using a Plasmon Enhanced Biosensor Platform

**DOI:** 10.1101/2020.09.02.20187070

**Authors:** Nathaniel C. Cady, Natalya Tokranova, Armond Minor, Nima Nikvand, Klemen Strle, William T. Lee, William Page, Ernest Guignon, Arturo Pilar, George N. Gibson

**Author notes:** Corresponding Author: Nathaniel C. Cady, PhD SUNY Polytechnic Institute.

## Abstract

The 2019 SARS CoV-2 (COVID-19) pandemic has highlighted the need for rapid and accurate tests to diagnose acute infection and determine immune response to infection. In this work, a multiplexed grating-coupled fluorescent plasmonics (GC-FP) biosensing approach was shown to have 100% selectivity and sensitivity (n = 23) when measuring serum IgG levels against three COVID-19 antigens (spike S1, spike S1S2, and the nucleocapsid protein). The entire biosensing procedure takes less than 30 min, making it highly competitive with well-established ELISA and immunofluorescence assays. GC-FP is quantitative over a large dynamic range, providing a linear response for serum titers ranging from 1:25 to 1:1,600, and shows high correlation with both ELISA and a Luminex-based microsphere immunoassay (MIA) (Pearson r > 0.9). Compatibility testing with dried blood spot samples (n = 63) demonstrated 100% selectivity and 86.7% sensitivity. A machine learning (ML) model was trained to classify dried blood spot samples for prior COVID-19 infection status, based on the combined antibody response to S1, S1S2, and Nuc antigens. The ML model yielded 100% selectivity and 80% sensitivity and demonstrated a higher stringency than a single antibody-antigen response. The biosensor platform is flexible and will readily accommodate detection of multiple immunoglobulin isotypes. Further, it uses sub-nanogram quantities of capture ligand and is thus readily modified to include additional antigens, which is shown by the addition of RBD in later iterations of the test. The combination of rapid, multiplexed, and quantitative detection for both blood serum and dried blood spot samples makes GC-FP an attractive biosensor platform for COVID-19 antibody testing.

## 1. INTRODUCTION

The rapid spread of the 2019 SARS CoV-2 (COVID-19) pandemic has established an urgent need for accurate diagnostic technologies (Pascarella et al. 2020). The majority of testing performed to date has utilized reverse transcription polymerase chain reaction (RT-PCR) for identifying viral genomic RNA. This high-throughput approach enables accurate diagnosis of acute infection but does not provide information about the immune responses of infected individuals. Due to the wide range in severity of this disease, many individuals remain asymptomatic or have mild symptoms, defining a population that is not tested at the time of acute infection (Pascarella et al. 2020). For these patients, the immune response to past COVID-19 infection is the best measure of exposure, making serologic testing critical. Such tests are also needed to confirm the presence or absence of protective immunity for individuals, especially as vaccines are developed and administered. Immune response to COVID-19 infection is variable, and the corresponding serological response (measured by antibody/immunoglobulin titer) may be linked to disease symptom severity, length of infection, and multiple patient-specific factors (Sethuraman et al. 2020; To et al. 2020). For these reasons, quantitative detection of the antibody response to COVID-19 is critical to our response to this pandemic.

To assess the antibody response to COVID-19 infection, a number of serological testing methods have been developed. Most of these tests focus on detection of antibodies, including immunoglobulin G (IgG) and/or immunoglobulin M (IgM) raised against viral antigens. These tests are typically performed using whole blood, blood serum, or blood plasma. Most testing methods utilize recombinant viral antigens as the target, and assess immunoglobulin binding to these antigens using a variety of direct and indirect detection methods. The most widely used approach is the enzyme linked immunosorbent assay (ELISA) (Amanat et al. 2020; Karp et al. 2020; Randad et al. 2020). ELISA-based testing enables high throughput (processing many samples in parallel), but is typically limited to a single antigen per well (Infantino et al. 2020; Younes et al. 2020). Thus, accurate diagnosis with this approach requires a robust antibody response against a single antigen, unless samples are tested in replicate, increasing cost, complexity and time to result. Multiplexed testing enables detection of immunoglobulin binding to more than one antigen within a single tube, well, plate or slide. These include, but are not limited to MIAs such as Luminex-based assays (Ayouba et al. 2020; Randad et al. 2020), fluorescent protein microarrays (Hedde et al. 2020), and direct/label-free array technologies (Steiner et al. 2020). The key challenges with these approaches are achieving high sensitivity, and low rate of false positives (selectivity), and quantitative measurement of immunoglobulin concentration/titer across a large dynamic range. Further, while these approaches increase assay content, they have a commensurate increase in assay cost.

Sample collection is a key challenge with implementing immunological / serological testing for large populations. For serology, blood samples are typically obtained by venipuncture, followed by blood plasma or serum preparation. This requires test subjects to travel to a medical clinic or blood collection site, which is burdensome for the participant and requires a complex chain of custody and/or cold chain for samples. Isolation of plasma requires additional processing steps and dedicated equipment. Alternatively, a simple finger stick and dried blood spotting allows self-collection, minimizing effort and likely increasing participation (Au – Grüner et al. 2015; Malsagova et al. 2020; Thevis et al. 2020; Vázquez-Morón et al. 2019). Samples collected in this manner may be maintained at ambient temperature and can be shipped using mail or courier service without the need for refrigeration or post-collection processing. Dried blood spots (DBS) have been successfully utilized for immunological/serological testing for multiple viral diseases, including hepatitis C, HIV, and COVID-19 (Karp et al. 2020; Malsagova et al. 2020; Vázquez-Morón et al. 2019).

Previously, we reported an enhanced fluorescence biosensor for multiplexed detection of antibodies for Lyme disease diagnosis (Chou et al. 2020; Chou et al. 2019; Chou et al. 2018). In this approach, gold-coated nanoscale grating surfaces were modified with target antigens in a microarray format and then used to detect IgG or IgM binding from blood serum or plasma. Surface plasmons generated during illumination of the gold-coated biosensor chip actively enhance fluorescence emission intensity, yielding a high-sensitivity fluorescence detection platform. We have termed this approach “grating-coupled fluorescent plasmonics” (GC-FP). To achieve high sensitivity and an added measure of selectivity, fluorophore-tagged antibodies (against IgM or IgG) are applied during a secondary labeling step. The entire detection process can be completed in less than 30 minutes with high sensitivity and specificity.

In this work we have used the GC-FP biosensor platform to develop a rapid immunoassay for simultaneous detection of antibodies against three COVID-19 spike protein antigens (RBD, S1, and S1S2) and the COVID-19 nucleocapsid protein (Nuc). Using serum, we have achieved 100% specificity and sensitivity for diagnosing prior COVID-19 infection, and for DBS we have demonstrated 100% specificity and sensitivity as high as 86.9%. For serum samples, GC-FP results are highly correlated with established testing methods (ELISA and MIA) and can be used to detect IgG, IgM and IgA antibody levels. The assay also has a large linear dynamic range across multiple orders of concentration, making it possible to determine antibody titer. Because antibody titer against COVID-19 antigens has been positively correlated with viral neutralization capacity (To et al. 2020), our test therefore has the potential to reveal the level of a subject’s immune response. Taken together, GC-FP and DBS may address key limitations in current serological testing by facilitating rapid, inexpensive, quantitative, and high-content serologic assays suitable for widespread testing of individuals without the need for clinical office visits.

## 2. MATERIALS AND METHODS

### 2.1 Materials

COVID-19 antigens including the full-length nucleocapsid protein (Nuc), the S1 fragment of the spike protein (S1), the full-length extracellular domain of the spike protein (S1S2), and the receptor binding domain of the spike protein (RBD) were all obtained from Sino Biological, Inc. Also obtained from Sino Biological, Inc. were human serum albumin (HSA), the S1 domain of the 2005 SARS coronavirus spike protein (WH20 isolate, abbreviated “SARS-S1”), and human Influenza B nucleoprotein (B/Florida/4/2006 isolate, abbreviated “Flu Nuc”). An additional positive control protein, human IgG protein (Hum IgG), was obtained from ThermoFisher Scientific. SuperBlock blocking buffer and phosphate buffered saline (PBS) were obtained from ThermoFisher Scientific, while TWEEN-20 solution was obtained from Sigma Aldrich. PBS-TWEEN (PBS-T) solution consisting of PBS + 0.05% v/v TWEEN-20 was prepared on a daily basis for all experiments. Alexa Fluor 647 labeled anti-human IgG (heavy and light chain) and anti-human IgM (heavy chain) were obtained from Invitrogen/ThermoFisher Scientific. Alexa Fluor 647 labeled anti-human IgA was obtained from Southern Biotech. ELISA testing was performed using COVID-19 human IgG testing kits from RayBiotech for full-length COVID-19 nucleocapsid protein (Nuc) and the S1 domain of the spike protein. The manufacturer’s protocol was followed for performing these ELISA assays.

### 2.2 Grating-Coupled Fluorescent Plasmonic (GC-FP) Biosensor Chip Preparation

Gold coated grating-coupled fluorescent plasmonic (GC-FP) biosensor chips were fabricated at SUNY Polytechnic Institute’s Center for Semiconductor Research using standard photolithographic patterning, etching and metal deposition techniques, as described previously (Chou et al. 2020; Chou et al. 2019). Double-sided adhesive gaskets and acrylic or polycarbonate fluidic covers for the GC-FP biosensor chips were either fabricated at SUNY Polytechnic Institute or obtained from Ciencia, Inc.

For use in the COVID-19 detection assay, GC-FP chips were printed with an array of approximately 400 μm diameter spots of target and control antigens/proteins using an ArrayIt SpotBot II microarray printer. All printed proteins/antigens were first diluted to 500 μg/μl in phosphate buffered saline (PBS) and then further diluted 1:1 just prior to printing with GBL protein array printing buffer (Grace Bio-Labs) All proteins/antigens used for printing were kept at –20 °C at 500 μg/μl for long-term storage. Target antigen spots were printed in triplicate, while negative and positive controls were printed in duplicate or triplicate on each GC-FP biosensor chip. After printing, GC-FP biosensor chips were allowed to dry in ambient conditions for a minimum of 30 min, then placed in a desiccant-filled box for long term storage.

### 2.3 Biological Samples

Human blood serum samples were obtained from donors within New York state or from the Wadsworth Center, New York State Department of Health. Negative samples were collected prior to the 2019 SARS CoV-2 pandemic. Positive samples were convalescent serum specimens received as part of the Wadsworth Center’s Diagnostic Immunology Laboratory (DIL) testing program that were SARS-CoV-2 confirmed by RT-PCR and at least 21 days post symptom onset. Samples were pre-screened by the DIL using a multiplex MIA with both the CoV-2 nucleocapsid (Nuc) protein and the 2019 SARS CoV-2 receptor binding domain (RBD) on a Luminex detection platform(Yang et al. 2020). The specimens were defined as reactive if one or both of the antigens was reactive. To determine reactive vs. non-reactive, an MIA assay cut-off was determined using 94 normal serum samples (collected prior to 2019). Cut-off values were based on the mean of the medial fluorescence intensity (MFI) plus 6 standard deviations. Samples received from the DIL for GC-FP analysis were tested blind (no sample information provided). After GC-FP test results were obtained, DIL testing results were provided for the purpose of comparison.

Additional negative control samples (collected prior to 2019) were obtained from healthy individuals through the Lyme Disease Biobank, and were used previously as negative controls for Lyme disease testing (Chou et al. 2020). Blood serum was prepared after venipuncture and collection in serum collection tubes by centrifugation at 1,000 rpm for 15 min, followed by removal of the supernatant (serum). All serum samples were kept at –20 °C for short-term storage, or –80 °C for long-term storage.

In addition to blood serum, blood samples were also collected by finger stick / dried blood spotting. 27-gauge lancet devices and Whatman 903 protein saver collection cards were sent to volunteers with instructions. Blood sampling and testing was approved by the SUNY

Polytechnic Institute Institutional Review Board (protocol #IRB-2020–10). Blood droplets were collected, allowed to dry, and then either hand delivered or mailed (via US Postal Service) to SUNY Polytechnic Institute. Following receipt of DBS samples, a sterile 6 mm diameter biopsy punch was used to remove samples from the collection cards. These disks were then soaked in 500 μl of PBS-T solution overnight (approximately 12 hr.) at 4 °C with gentle rocking to liberate blood / serum proteins, including immunoglobulins. Samples were then analyzed using GC-FP biosensor chips within 24 hr. of being processed.

### 2.4 GC-FP Detection Assay

Prior to performing GC-FP detection assays, GC-FP chips were assembled with an adhesive gasket and flow cell cover, filled with SuperBlock blocking buffer to prevent non-specific surface interactions, then incubated at room temperature for a minimum of 15 min. Following assembly and blocking, GC-FP chips were placed in a custom fluidic apparatus to provide sequential flow of sample and reagents. Using this setup, the following steps were followed for all samples: 1) 500 μl of PBS-T at 100 μl/min, 2) 400 μl of diluted human blood serum or extracted dried blood spot sample at 50 μl/min, 3) 500 μl of PBS-T at 100 μl/min, 4) 400 μl of Alexa 647 anti-human IgG / IgM (diluted 1:400 in PBS-T) at 100 μl/min, and 5) 500 μl of PBS-T at 100 μl/min. GC-FP chips were then analyzed in a customized Ciencia, Inc. fluorescent plasmonic imaging instrument. For serum testing, a standard dilution of serum in PBS-T (1:25) was used. For dried blood spot testing, undiluted extract from the 6 mm diameter segment of the blood collection card was used. Ciencia image analysis LabView software was then used to define a region of interest (ROI) for each individual spot on the GC-FP biosensor chip and the fluorescence intensity of each spot was measured. The fluorescence intensity of all spots was normalized to the human IgG (Hum IgG) internal control spots on each chip, to account for variability between individual chips and individual experiments. For normalization, the Hum IgG spots on each chip were set to a baseline intensity of 100 (arbitrary units) and all remaining spots were adjusted accordingly.

### 2.5 Data Analysis

Normalized spot intensity data was exported from the software and further analyzed using Microsoft Excel and/or GraphPad Prism 8.0 software. The average and standard deviation of normalized intensity values were calculated for all replicate protein/antigen spots, including COVID-19 target antigens (S1, S1S2, Nuc, RBD), positive controls (Flu Nuc, Hum IgG), and negative controls (HSA, SARS S1). To further account for variation between chips and experiments, normalized intensity data for positive control and COVID-19 antigen spots (mean intensity, 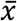) were divided by the average negative control spot intensity, plus three times the standard deviation (σ) of the negative control spot intensity 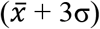 to produce a detection metric as follows:

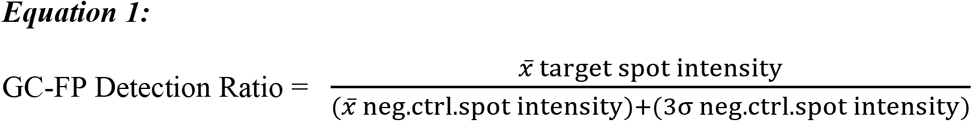

To determine the threshold detection ratio values for diagnosis, serum or dried blood spots samples with confirmed COVID-19 history (confirmed positive or confirmed negative/no known exposure) were used to perform receiver operator characteristic (ROC) analysis using GraphPad Prism 8.0. Detection ratios for each COVID-19 antigen, for both positive and negative samples, were entered into Prism 8.0 and analyzed with the ROC analysis package, with area under the curve (AUC) analysis performed for each ROC curve. The threshold detection ratio needed to obtain 100% specificity (with the MFI results as the gold standard) and maximize sensitivity (using a 95% confidence interval) was chosen for each ROC curve, and overall p-value was reported. GraphPad Prism 8.0 was used to perform an unpaired T-test on the calculated detection ratios for all COVID-19 antigens (for both serum and dried blood spots) to determine whether there was a significant difference in the mean GC-FP detection ratio for positive vs. negative samples. This software was also used to perform fitting, correlation, and regression analysis on quantitative GC-FP and ELISA test results. Linear fitting was used for GC-FP data, while partial least squares fitting was used for ELISA data. R-square analysis was also performed for all regressions, to indicate goodness of fit.

A support vector machine (SVM) based machine learning approach was used to analyze GC-FP detection data, and was implemented with previously described, freely available SVM software (LibSVM - http://www.csie.ntu.edu.tw/~cjlin/libsvm) (Chang and Lin 2011). The nu-SVC package within LibSVM was utilized with sigmoid kernel, and a grid search for cost and gamma parameters was conducted to maximize the prediction accuracy of the SVM model.

## 3. RESULTS AND DISCUSSION

### 3.1 Detection Assay Development and Characterization

Rapid (less than 30 min), multiplexed detection of immunoglobulin binding to COVID-19 antigens was performed using our previously described GC-FP biosensing approach and a Ciencia, Inc. fluorescent plasmonic imaging instrument (Chou et al. 2020; Chou et al. 2019; Chou et al. 2018). By using commercially available, recombinant COVID-19 specific antigens, GC-FP microarrays were readily produced using standard protein spotting techniques. An example GC-FP microchip for COVID-19 immunological analysis is shown in Figure 1A, and a conceptual diagram of the assay is shown in Figure 1B. Using this approach, COVID-19 specific antigens and control proteins were immobilized on the GC-FP biosensor chips in a variety of configurations (Supplementary Figure S1).

**Figure 1.**
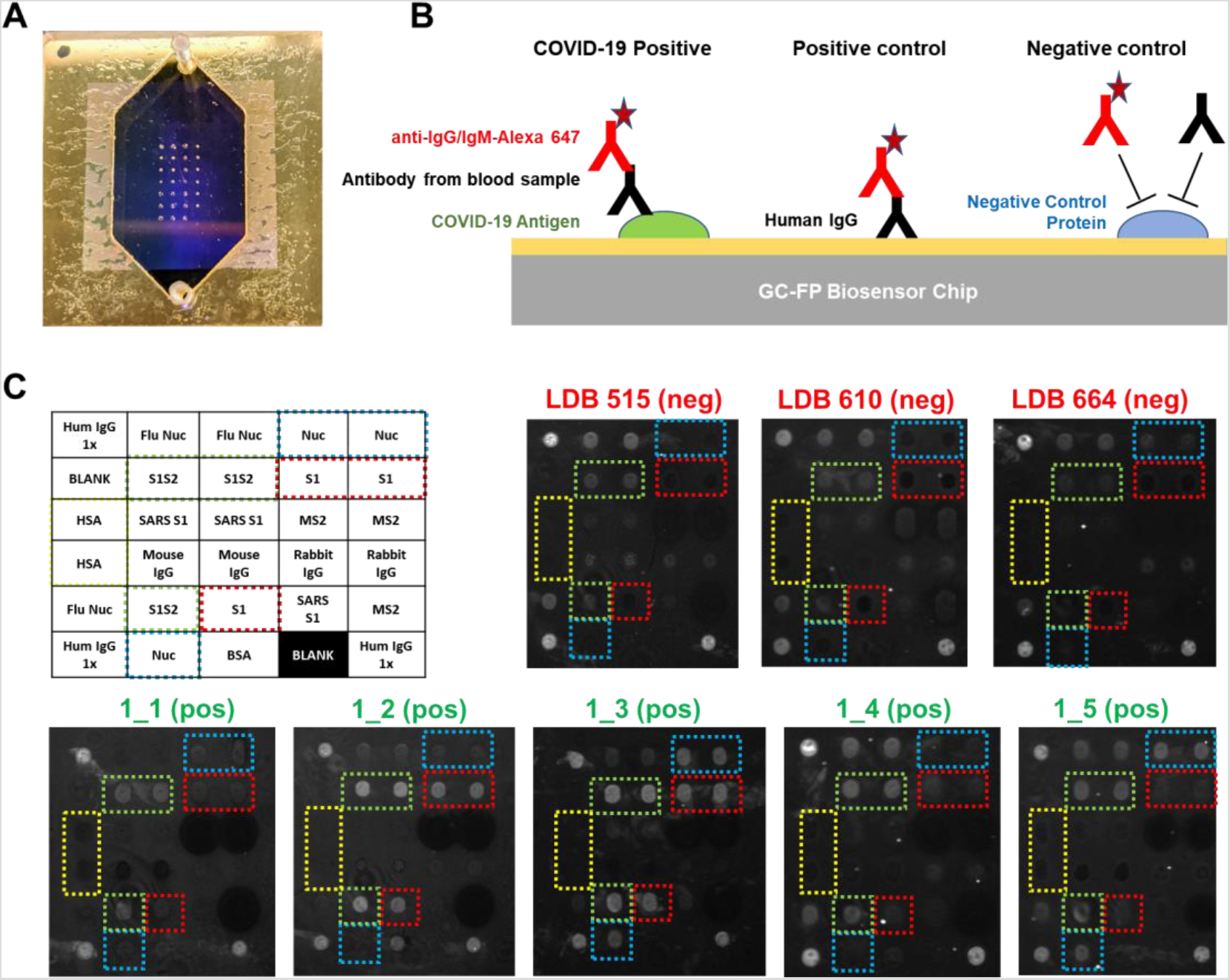
A) GC-FP biosensor chip shown with gasket and fluidic cover attached. The array of antigen/protein spots can be seen imprinted on the gold surface of the chip. B) Schematic of the GC-FP COVID-19 assay approach. COVID-19 antigens or control proteins were spotted onto GC-FP biosensor chips, then assessed for antibody binding from human blood samples. Subsequent labeling with Alexa Fluor 647 tagged anti-human IgG was used for the enhanced fluorescence detection output. C) Enhanced fluorescence images of GC-FP biosensor chips (v1) processed with negative control serum (blood serum collected >2 years prior to the COVID-19 pandemic and serum from subjects who were >2 weeks convalescent from PCR-confirmed COVID-19 infection. Boxes with dotted outlines indicate paired spots of key COVID-19/SARS CoV-2 antigens, S1, S1S2, and Nuc.

Initial testing utilized serum samples from COVID-19 convalescent, previously RT-PCR positive, subjects who were expected to have an antibody response to the COVID-19 antigens included in our assay. All subjects had fully recovered from infection and were more than 2 weeks convalescent. As negative controls, serum samples from healthy individuals collected prior to 2019 were used. These samples had been previously used in our Lyme disease diagnostic development and were expected to show little to no IgM or IgG response to COVID-19 antigens (Chou et al. 2020). Shown in Figure 1C, negative control samples showed no GC-FP response for IgG binding to COVID-19 Nuc and S1 antigens, and in some cases, very weak response for full-length spike S1S2 extracellular domain antigen. These chips (v1) did not include RBD antigen, and therefore IgG response to this antigen was not measured. Similar experiments were performed to determine the IgM response for these samples, with equivalent results (data not shown). Due to the generally higher specificity of IgG, we determined that IgG detection alone was suitable for all follow-on testing.

### 3.2 GC-FP Antibody Detection in Human Serum and Dried Blood Spot Samples

Following initial testing with v1 GC-FP chips, testing was performed using v2 – v4 chips that reduced the number of negative control proteins, and increased the number of replicates for positive (Hum IgG) and negative (HSA) control spots. Using these arrays, 23 different human blood serum samples, and 24 dried blood spot samples (with verified infection status) were tested. All positive samples were previously RT-PCR positive for COVID-19 infection, and negative samples were either RT-PCR negative for COVID-19 infection, or from individuals who had no known exposure, nor symptoms of COVID-19 infection. Samples were tested at a dilution rate of 1 part serum to 25 parts PBS-T (for serum) or undiluted (for dried blood spot extracts). Raw GC-FP fluorescence intensity data were normalized as described in the Materials and Methods section, and a “GC-FP detection ratio” was calculated to account for chip-to-chip differences and any variability in processing conditions. As described, the GC-FP detection ratio compares the normalized average GC-FP intensity for target antigens against the average (plus 3σ) negative control intensity. Thus, the reported GC-FP detection ratio provides a measure of signal above background / non-specific binding signal, while also accounting for spot-to-spot variability. The results of these experiments are shown in Figure 2 and in Tables 1 and Supplementary Table T1. The average GC-FP detection ratio for COVID-19 positive samples vs. COVID-19 negative samples was significant for all antigens, for both serum and dried blood spot samples (Mann-Whitney U-test, p < 0.05).

**Figure 2.**
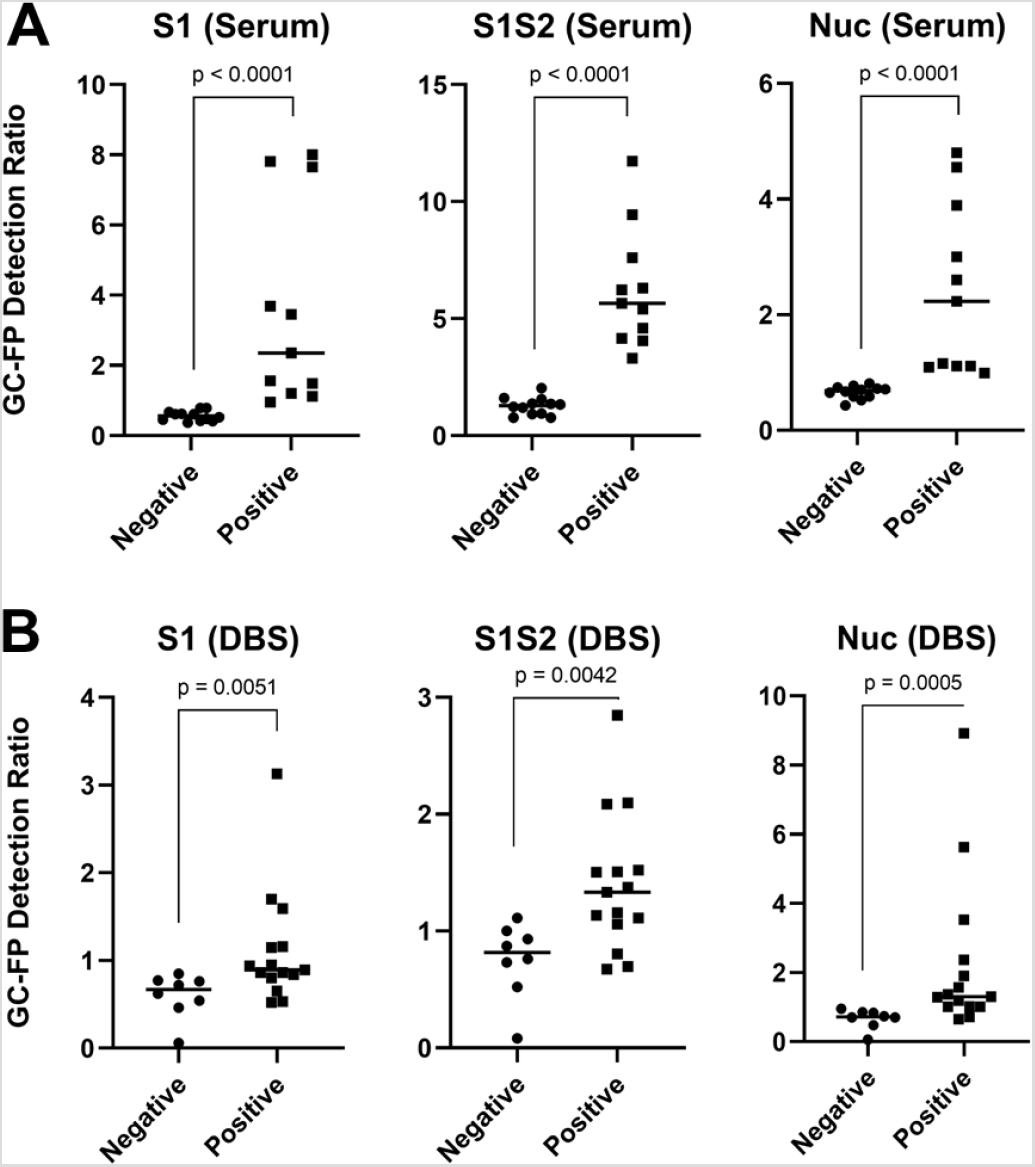
GC-FP assay results for serum (A) and dried blood spot (B) samples from subjects with verified positive COVID-19 infection status. GC-FP detection ratio is plotted for each individual antigen. A significant difference between the average positive and negative GC-FP detection ratios (horizontal line in each plot) was confirmed using the Mann-Whitney U-test, with corresponding p values shown in each plot. ELISA (450 nm absorbance values) for the COVID-19 nucleocapsid protein were compared to GC-FP detection ratio results for a subset of serum samples (C), while Luminex MIA results (MFI values) were compared to GC-FP detection ratio results for a second subset of samples (D). Pearson correlation coefficients (r) and R-squared values from correlation are shown, with associated p-values. Dashed lines indicate linear regression fit to the data, while dotted lines indicate the 95% confidence intervals for correlation analysis.

**Table 1.** GC-FP detection ratio data for serum and dried blood spot samples with verified infection status. ELISA and Luminex MIA data are shown for serum samples tested with these assays. Samples scored positive (GC-FP detection ratio above the ROC threshold) are denoted by bold text. The Nuc diagnostic score and machine learning (ML) score are shown for dried blood spot samples. Notes regarding each sample are shown in the last column.

| Serum Samples |  |  |  |  |  |  |
| --- | --- | --- | --- | --- | --- | --- |
| Sample ID | S1 | S1S2 | Nuc | Nuc-ELISA<br>(score / A450) | Nuc-MIA<br>(score / MFI) | Notes |
| 2_1 | 0.67 | 1.55 | 0.81 | neg / 0.054 | n/a | no known exposure |
| 2_2 | 0.61 | 0.94 | 0.72 | neg / 0.027 | n/a | no known exposure |
| 2_3 | 0.79 | 0.77 | 0.74 | neg / 0.013 | n/a | no known exposure |
| 2_4 | 0.79 | 0.77 | 0.67 | neg / 0.046 | n/a | no known exposure |
| 2_5 | 0.52 | 1.33 | 0.77 | neg / 0.032 | n/a | no known exposure |
| 2_6 | 0.61 | 1.60 | 0.70 | neg / 0.031 | n/a | no known exposure |
| 2_7 | 0.62 | 1.37 | 0.71 | neg / 0.037 | n/a | no known exposure |
| DIL0526-26 | 0.42 | 0.92 | 0.59 | n/a | neg / 259 | 2009 pre-COVID |
| DIL0526-27 | 0.41 | 1.20 | 0.52 | n/a | neg / 421 | 2009 pre-COVID |
| DIL0526-28 | 0.45 | 1.23 | 0.65 | n/a | neg / 1,563 | 2009 pre-COVID |
| DIL0526-29 | 0.48 | 2.03 | 0.59 | n/a | neg / 238 | 2009 pre-COVID |
| DIL0526-30 | 0.37 | 1.36 | 0.43 | n/a | neg / 719 | 2009 pre-COVID |
| 1_1 | **1.20** | **3.31** | **1.16** | **pos / 0.123** | n/a | Positive by RT-PCR |
| 1_2 | **3.69** | **5.65** | **1.09** | **pos / 0.178** | n/a | Positive by RT-PCR |
| 1_3 | **7.65** | **11.73** | **4.80** | **pos / 1.577** | n/a | Positive by RT-PCR |
| 1_4 | **1.49** | **5.41** | **0.99** | **neg / 0.043** | n/a | Positive by RT-PCR |
| 1_5 | **1.56** | **4.59** | **2.23** | **pos / 0.491** | n/a | Positive by RT-PCR |
| DIL0526-3 | **7.81** | **9.44** | **3.89** | n/a | **pos / 45,543** | Positive by RT-PCR |
| DIL0526-5 | **8.00** | **7.61** | **4.55** | n/a | **pos / 47,301** | Positive by RT-PCR |
| DIL0526-8 | **3.45** | **6.23** | **3.00** | n/a | **pos / 39,255** | Positive by RT-PCR |
| DIL0526-13 | **2.35** | **4.06** | **1.11** | n/a | **pos / 11,577** | Positive by RT-PCR |
| DIL0526-18 | **1.12** | **6.31** | **2.60** | n/a | **pos / 34,715** | Positive by RT-PCR |
| DIL0526-21 | **0.95** | **4.16** | **1.11** | n/a | **pos / 28,806** | Positive by RT-PCR |
| Dried Blood Spot Samples (Verified Infection Status) |  |  |  |  |  |  |
| Sample ID | S1 | S1S2 | Nuc | Nuc Score | ML Score | Notes |
| COV_5 | 0.54 | 0.73 | 0.70 | neg | neg | Negative by RT-PCR |
| COV_6 | 0.72 | 1.11 | 0.70 | neg | neg | Negative by RT-PCR |
| COV_7 | 0.06 | 0.08 | 0.06 | neg | neg | Negative by RT-PCR |
| COV_9 | 0.85 | 1.00 | 0.95 | neg | neg | Negative by RT-PCR |
| COV_14 | 0.46 | 0.52 | 0.48 | neg | neg | Negative by RT-PCR |
| COV_16 | 0.77 | 0.93 | 0.84 | neg | neg | Negative by RT-PCR |
| COV_18 | 0.62 | 0.76 | 0.74 | neg | neg | Negative by RT-PCR |
| COV_12 | 0.76 | 0.87 | 0.85 | neg | neg | IgG and IgM negative |
| COV_2 | 0.65 | 0.80 | 0.72 | neg | neg | Positive by RT-PCR |
| COV_3 | **1.59** | **1.13** | **5.63** | *pos* | *pos* | Positive by RT-PCR |
| COV_13 | 0.53 | 0.67 | 0.65 | neg | neg | Positive by RT-PCR |
| COV_17 | **0.94** | 1.11 | **1.01** | *pos* | *pos* | Positive by RT-PCR |
| COV_28 | 0.80 | **1.37** | **3.53** | *pos* | *pos* | Positive by RT-PCR |
| COV_29 | **1.70** | **1.51** | **1.57** | *pos* | *pos* | Positive by RT-PCR |
| COV_40 | **1.15** | **1.50** | **2.37** | *pos* | *pos* | Positive by RT-PCR |
| COV_41 | **0.86** | 1.06 | **1.02** | *pos* | *pos* | Positive by RT-PCR |
| COV_43 | 0.52 | 0.70 | **1.19** | *pos* | neg | Positive by RT-PCR |
| COV_44 | **3.13** | **2.85** | **8.92** | *pos* | *pos* | Positive by RT-PCR |
| NYC_1 | **1.16** | **2.09** | **1.29** | *pos* | *pos* | Positive by RT-PCR |
| NYC_5 | **0.95** | **2.09** | **1.90** | *pos* | *pos* | Positive by RT-PCR |
| COV_19 | **0.89** | **1.52** | **1.37** | *pos* | *pos* | IgG pos., no PCR test |
| COV_38 | **0.86** | **1.33** | **1.30** | *pos* | *pos* | IgG pos., no PCR test |
| COV_24 | 0.84 | **1.16** | **1.01** | *pos* | *pos* | IgM pos., no PCR test |

GC-FP detection ratios for serum samples were compared to an ELISA-based assay and a Luminex-based MIA (Yang et al. 2020), when using COVID-19 nucleocapsid (Nuc) as the target antigen. As shown in Figure 2C and Figure 2D, the measured GC-FP detection ratios for serum samples are highly correlated with both ELISA absorbance values (Pearson r = 0.944, R-squared = 0.892) and MIA fluorescence intensity (MFI) values (Pearson r = 0.939, R-squared = 0.882). These results demonstrate that the GC-FP detection approach provides comparable immunodetection results to established, gold-standard methods.

To determine if individual IgG/antigen responses could be used for diagnostic purposes, receiver operator characteristic (ROC) analysis was performed (Supplementary Figure S2). For serum samples, 100% sensitivity and specificity could be achieved when the following GC-FP detection ratio thresholds were met: S1 = 0.87, S1S2 = 2.67, Nuc = 0.9. Area under the curve (AUC) analysis yielded values of 1.0 for all antigens (p < 0.0001) indicating extremely high confidence in diagnosis using individual antigens, when GC-FP detection ratios met or exceeded the ROC thresholds. For dried blood spot samples, ROC analysis yielded 100% specificity and variable sensitivity when the following GC-FP detection ratio thresholds were met: S1 = 0.855, S1S2 = 1.12, and Nuc = 0.98. AUC values ranged from 0.85 to 0.92, all of which are well above the cut-off value of 0.7 that we have successfully used for GC-FP based Lyme disease diagnosis (Chou et al. 2020). When these GC-FP detection ratio thresholds were exceeded (to maintain 100% specificity) assay sensitivity was relatively low for S1 and S1S2 antigens (66.7% for both) but increased to 86.7% for the nucleocapsid antigen (Nuc). Table 1 lists GC-FP testing results from all dried blood spot samples.

The observed reduction in sensitivity for DBS vs. serum samples could be due to the sample format, especially since sample stability and extraction are more variable for dried blood spots vs. blood serum. Other reasons for the reduction in sensitivity could be variability in individual immune responses and/or reporting consistency for the research subjects who provided samples. Research subjects self-reported prior COVID-19 test results (RT-PCR and/or clinical antibody tests), and copies of clinical test results were not provided (as per our approved IRB protocol). Thus, prior infection status could not be independently verified. Other studies have shown that individual antibody responses are variable (Amanat et al. 2020), which would also affect assay sensitivity. For example, subjects COV_2 and COV_13 were both previously positive via RT-PCR, but GC-FP testing showed no detectable antibody signal against S1, S1S2, or Nuc (Table 1) from their dried blood spot samples. While this could be due to lack of sensitivity of the assay itself, it could also be due to poor immune response and/or low antibody production in response to infection. Importantly, no false positives were observed within the limited number of samples that were tested. To definitively validate the sensitivity and specificity of the assay, a much larger cohort of samples will be required.

In addition to testing samples with known COVID-19 infection history, 39 additional dried blood spot samples were received and tested. For these samples, information was provided about exposure to infected individuals, potential disease symptoms, or complete lack of exposure, but none of the subjects had been tested with a COVID-19 RT-PCR test. The results from GC-FP testing for these samples are shown in Supplementary Table T1. Due to the fact that COVID-19 infection status was unverified for these samples, sensitivity and specificity could not be determined.

### 3.3 Multiplexed Data Analysis

One advantage of the GC-FP approach is that antibody binding information is collected simultaneously for multiple antigens (e.g. RBD, S1, S1S2, and Nuc). Scoring samples based on a composite antibody response (to all antigens) could provide increased diagnostic accuracy and a more complete understanding of a subject’s antibody response to infection. A support vector machine (SVM) based machine learning (ML) approach was used to differentiate and classify samples based on their antibody response to three target antigens that were used for all samples tested (S1, S1S2, and Nuc). ML approaches have been used extensively for classification and diagnosis when data from multiple biomarkers or targets is available (Sarkar and Saha 2019; Uddin et al. 2019). The SVM approach to ML involves supervised learning, where a well characterized (labeled) data set is used to train the model, and that model is then used to classify unlabeled data. An established SVM software (LibSVM) (Chang and Lin 2011) was trained using GC-FP data from serum samples (Table 1). GC-FP serum testing results had yielded 100% sensitivity and specificity (for all antigens used) with ROC analysis, and when detection ratios were plotted in a 3D plot (Figure 3A) positive and negative samples were well-separated. Taken together, these analyses established serum data as an ideal training set for the ML model. After training, the ML model was challenged with 10-fold cross-validation on unlabeled serum data, yielding 100% selectivity and sensitivity, which matched ROC analysis for individual antigens. After training and validation, DBS data (from subjects with verified prior COVID-19 status and those with unverified status) were classified with the SVM model (Figure 3B & 3C, Table 1, Supplementary Table T1). For dried blood spots with verified prior infection status, the SVM model classified samples with 80% sensitivity and 100% selectivity. For dried blood spots with unverified prior infection status, SVM classification resulted in better correlation with presumed infection status than when scoring with individual antibody responses (S1, S1S2, or Nuc).

**Figure 3.**
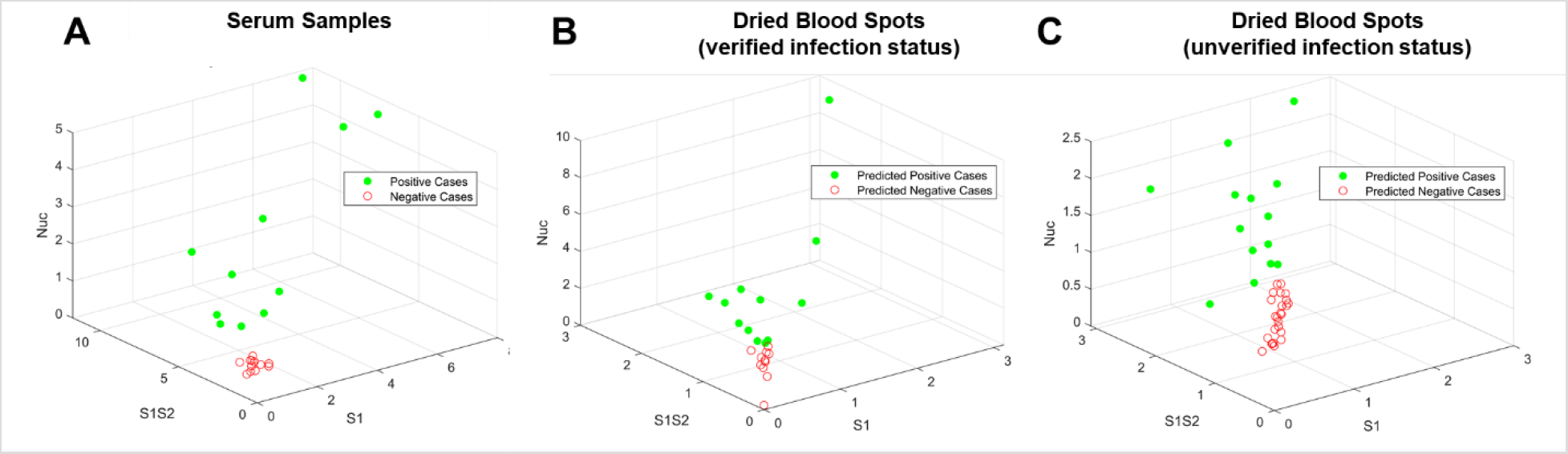
Visualization of testing data following machine learning-based analysis. (A) The GC-FP detection ratio data for all serum samples were plotted as a function of the three antigens S1, S1S2, and Nuc. These data were then used to train an SVM ML model, which was used to classify dried blood spot sample data from (B) subjects with verified COVID-19 infection status, and (C) subjects with unverified COVID-19 infection status.

The ML sensitivity (80%) represents a slight reduction in sensitivity obtained when using only the Nuc antigen (86.7%) for scoring. This was due to a scoring difference for subject COV_43, who previously tested positive for COVID-19 by RT-PCR. This subject’s Nuc antibody response (1.19) was above the ROC-determined detection threshold (0.98), but S1 and S1S2 were well below the ROC-determined thresholds. The ML model classified this sample as negative, which is an indication that the model is more stringent than using a single antibody-antigen interaction for scoring.

Interestingly, the ML model classified three samples from subjects with no known COVID-19 exposure as positive, while scoring with only Nuc resulted in five such samples scoring positive. Prior infection status was not verified for these samples, thus it is not clear if these results are actually false positives. If these are indeed false positives, the ML based classification provides better selectivity vs. scoring with only the Nuc antibody response. The ML based classification also scored a subject (NYC_6) as negative, while scoring with just the Nuc antigen resulted in a positive diagnosis (Supplementary Table T1). This subject reported living with a COVID-19 positive individual, but never experienced disease symptoms. Because this individual did not obtain a COVID-19 RT-PCR test, it is unknown whether they were infected and asymptomatic, or never infected. Regardless, this example further illustrates the increased stringency of the ML based classification over single antigen scoring. While many additional samples will be needed to fully train the ML model, and to understand the true selectivity and sensitivity of the GC-FP assay, we have illustrated the potential for ML based scoring antibody responses to multiple antigens. Training the ML classification model is highly adaptable as more sample data are collected, and thus we anticipate ongoing improvement in the model and its ability to correctly classify samples.

### 3.4 Quantification of Antibody Titer and Comparison to ELISA

To assess GC-FP for quantitative determination of antibody concentration (titer), individual GC-FP chips were processed using dilutions of a COVID-19 positive serum sample received from the NYS Department of Health (sample DIL0526–3). Sample dilutions ranged from 1:25 to 1:25,600 and GC-FP testing results were compared to ELISA against S1 and Nuc, using commercial ELISA kits (Ray Biotech). The results of this experiment, shown in Figure 4A, demonstrate that GC-FP could detect IgG at a minimum dilution of 1:1,600 and that the commercial ELISA kit could detect IgG to a minimum dilution of 1:6,400. Beyond these dilutions, the respective assays could no longer reliably distinguish IgG biding signal from background / negative controls.

**Figure 4.**
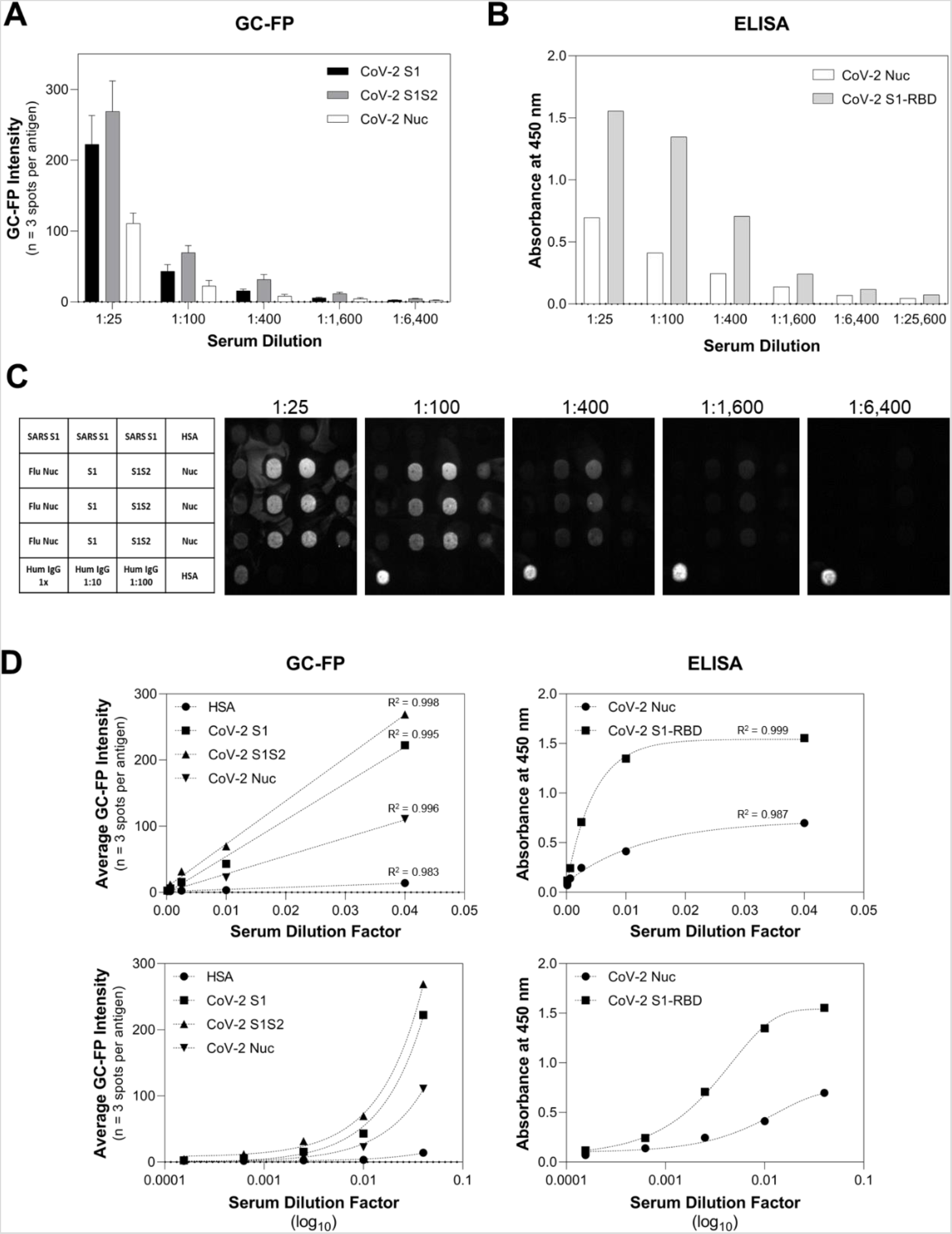
Quantitative comparison of GC-FP and ELISA for detection of IgG against multiple COVID-19 antigens. Sample DIL0526–3 was used for both GC-FP and ELISA testing, at dilutions ranging from 1:25 to 1:26,600 in PBS-T. GC-FP intensity is reported based on normalization to human IgG positive control signal, while ELISA data are reported as the resulting absorbance at 450 nm following the manufacturer’s recommended ELISA testing conditions.

When GC-FP and ELISA data were plotted as a function of dilution factor (Figure 4B) and fit with either linear regression (GC-FP) or partial least squares regression (ELISA), high goodness of fit (R-square > 0.98) was observed for all antigens. The GC-FP method exhibited a linear response across a large dynamic range (1:25 – 1:6,400 dilutions). The ELISA data retained goodness of fit across a larger range (1:25 – 1:25,600), but the response was nonlinear.

Recent COVID-19 antibody testing studies have shown that antibody titers (as determined by ELISA) of 1:80 – 1:160 are considered low, 1:320 as moderate, and >1:320 as high (Amanat et al. 2020; Wajnberg et al. 2020). Further, titers in the range of 1:320 or higher could be considered eligible for convalescent plasma donation (for convalescent plasma therapy)(Wajnberg et al. 2020). As shown in this work, GC-FP can detect antibodies down to 1:1,600 titer, and thus has the necessary sensitivity for determining clinically and therapeutically relevant seroconversion status. The fact that GC-FP has a linear response for all antigens tested makes it easier to quantify antibody concentrations across the full dynamic range, and to directly compare a subject’s response to different viral antigens.

### 3.5 GC-FP Detection of Multiple Immunoglobulin Isotypes

As a final experiment, three individual v4 GC-FP chips were processed with a COVID-19 positive, convalescent serum sample (from subject 1_3) and then assessed for binding by different antibody isotypes (IgG, IgM, and IgA), by labeling with Alexa Fluor 647 anti-human IgG, IgM, or IgA (Figure 5). A fourth chip was processed with dilution buffer (PBS-T) instead of serum, and then labeled with a mixture of Alexa 647 anti-human IgG, IgM, and IgA. Different antibody binding patterns were observed, based on the immunoglobulin isotype and the antigen being targeted (Figure 5B). IgG binding was highest for the Nuc antigen, IgM showed the highest binding to RBD, and IgA showed similar binding for S1, S1S2, and Nuc, but little binding to RBD.

**Figure 5.**
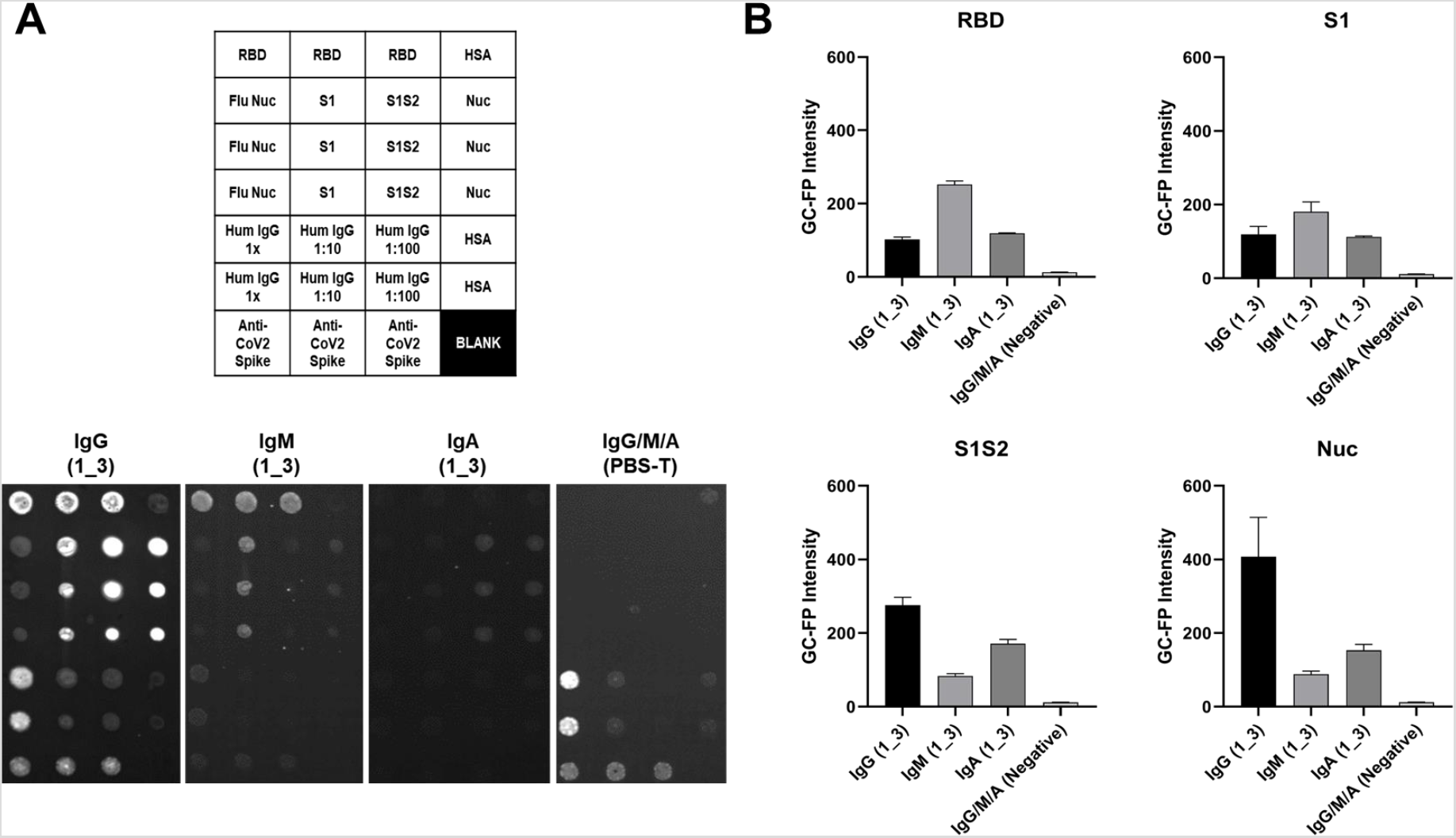
Serum sample 1_3 was tested on three separate v4 GC-FP biosensor chips that included RBD, S1, S1S2, and Nuc antigens. A fourth chip was processed using PBS-T as a negative control. The three chips tested with 1_3 serum were labeled with Alexa 647-tagged anti-human IgG, anti-human IgM, or anti-human IgA, while the negative control chip was labeled with a mixture of all three secondary antibodies. The mean GC-FP intensity (n = 3 spots per chip) is shown, for each of the COVID-19 antigens.

These results demonstrate the potential of the GC-FP approach to not only perform simultaneous, quantitative detection of antibody binding to multiple antigens, but also discrimination of antibody binding based on immunoglobulin isotype. As more samples are tested with the GC-FP platform, the major goal will be to retain 100% selectivity while maximizing sensitivity. One way to achieve this will be to compare antibody responses to additional COVID-19 antigens, as we have shown with the addition of RBD. Understanding antibody responses across various immunoglobulin classes may also be useful for determining the stage of an individual’s seroconversion response, and could be useful when analyzing other bodily fluids, such as saliva (Randad et al. 2020).

## 4. CONCLUSIONS

Rapid, accurate, and quantitative antibody tests are needed as part of the global response to the COVID-19 pandemic. In addition to epidemiological and seroconversion studies, such tests have the potential to confirm an individual’s immunity status following prior infection or vaccination. However, population studies have demonstrated considerable heterogeneity of the immune response between exposed individuals, as well as within a single individual over time.

The GC-FP assay developed in this work simultaneously measures antibody levels for multiple antigens, is quantitative across a large dynamic range, is sufficiently inexpensive to permit population-level studies, and yields a diagnostic result in less than 30 min. Compared to other methods, this represents a major reduction in assay time (compared to 2–3 hrs for competing approaches), without negatively impacting the accuracy or quantitative aspects of the test. Going beyond standard blood serum testing, this study also demonstrates that dried blood spot testing is a viable approach, that is both compatible with the GC-FP assay, and reduces the complexity of sample collection, handling and storage. Overall, this work provides justification for GC-FP as an alternative approach to serologic COVID-19 testing that overcomes key limitations of extant technologies.

## Data Availability

All data is available upon request.

## AUTHORS’ CONTRIBUTIONS

N.C.C. conducted experimental design, experimental work, data analysis, and was the primary author of this manuscript. N.T. and A.M. assisted with experimental work. G.G., E.G., W.P. and A.P. fabricated the GC-FP fluorescent imaging platform and wrote the analysis software used for testing. N.N. performed data analysis and machine learning analysis. K.S. assisted with writing, editing and data analysis, while W.L. provided serum samples and Luminex MIA data.

## DECLARATION OF COMPETING INTERESTS

A.P., E.G., and W.P. are all employees of Ciencia, Inc. and G.G. is a consultant for Ciencia, Inc.

## ACKNOWLEDGEMENTS

The authors thank Dr. Brion Winston and Capital Cardiology (Albany, NY) for assistance with collecting dried blood spot samples. We would also like to thank Kelly Howard, Mary Marchewka, and Karen Kulas for MIA specimen characterization. This work was supported by the State University of New York COVID-19 seed funding program.

## Notes

### Funding Statement

Funding was provided by the State University of New York.

### Author Declarations

Blood sampling and testing was approved by the SUNY Polytechnic Institute Institutional Review Board (protocol #IRB-2020-10).

### Summary of Updates

Addition of author.

